# Comparing the Performance of ChatGPT and GPT-4 versus a Cohort of Medical Students on an Official University of Toronto Undergraduate Medical Education Progress Test

**DOI:** 10.1101/2023.09.14.23295571

**Authors:** Christopher Meaney, Ryan S. Huang, Kevin (Jia Qi) Lu, Adam W. Fischer, Fok-Han Leung, Kulamakan Kulasegaram, Katina Tzanetos, Angela Punnett

## Abstract

**Background:** Large language model (LLM) based chatbots have recently received broad social uptake; demonstrating remarkable abilities in natural language understanding, natural language generation, dialogue, and logic/reasoning.

**Objective:** To compare the performance of two LLM-based chatbots, versus a cohort of medical students, on a University of Toronto undergraduate medical progress test.

**Methods:** We report the mean number of correct responses, stratified by year of training/education, for each cohort of undergraduate medical students. We report counts/percentages of correctly answered test questions for each of ChatGPT and GPT-4. We compare the performance of ChatGPT versus GPT-4 using McNemar’s test for dependent proportions. We compare whether the percentage of correctly answered test questions for ChatGPT or GPT-4 fall within/outside the confidence intervals for the mean number of correct responses for each of the cohorts of undergraduate medical education students.

**Results:** A total of N=1057 University of Toronto undergraduate medical students completed the progress test during the Fall-2022 and Winter-2023 semesters. Student performance improved with increased training/education levels: UME-Year1 mean=36.3%; UME-Year2 mean=44.1%; UME-Year3 mean=52.2%; UME-Year4 mean=58.5%. ChatGPT answered 68/100 (68.0%) questions correctly; whereas, GPT-4 answered 79/100 (79.0%) questions correctly. GPT-4 performance was statistically significantly greater than ChatGPT (P=0.034). GPT-4 performed at a level equivalent to the top performing undergraduate medical student (79/100 questions correctly answered).

**Conclusions:** This study adds to a growing body of literature demonstrating the remarkable performance of LLM-based chatbots on medical tests. GPT-4 performed at a level comparable to the best performing undergraduate medical student who attempted the progress test in 2022/2023. Future work will investigate the potential application of LLM-chatbots as tools for assisting learners/educators in medical education.

## 1 Introduction

Large language model (LLM) based chatbots (e.g. OpenAI ChatGPT and GPT-4, Google Bard, Facebook Llama, Anthropic AI Claude, and others) have demonstrated remarkable capabilities in various aspects of natural language understanding, natural language generation, dialogue and logic/reasoning [Bubeck et al., 2023].

In academic settings, these LLM-based chatbots have demonstrated human-like performance when applied across several heterogeneous high-stakes tests/exams (of varying difficulty levels, from various academic disciplines), which were often thought to require specialized training, knowledge, and reasoning skills [OpenAI, 2023].

In this study, we compare the performance of ChatGPT and GPT-4 against a cohort of medical students on an official University of Toronto undergraduate medical education progress test. Specifically, we compare the proportion of correct answers by medical students versus these LLM-based chatbots. We contextualize our findings in relation to a growing amount of literature applying LLM-based chatbots to high-stakes medical exams and discuss the application of LLM-based chatbots in medical education settings.

## 2 Methods

This cross-sectional study compared the performance of N=1057 medical students against two LLM-based chatbots (ChatGPT and GPT-4) on an official University of Toronto Undergraduate Medical Progress Test. The progress test was administered to students during the Fall-2022 and Winter-2023 semesters. The test consisted of 100 multiple-choice questions. The test was designed to assess medical knowledge and clinical decision making at the level of a graduating medical student and is administered to all medical students repeatedly during the course of training. The test is intended as a formative tool to support student learning in preparation for licensing examination and as a method of tracking their growing knowledge base. The test was based on the Medical Council of Canada examination objectives and covered the breadth of clinical presentation and diagnoses, population health and its determinants, and the legal, ethical and organizational aspects of medicine (https://mcc.ca/objectives/). Test items were developed by a trained team of item writers representing core medical disciplines with item review prior to and post-test administration.

The multiple-choice questions were entered into each of ChatGPT and GPT-4 exactly as written. The responses generated by each LLM chatbot were collected, and a single author (AP) scored whether the returned response was correct/incorrect. Additionally, information was collected regarding response generation time (seconds), response length (number of characters), whether a rationale for the generated response was provided (yes/no), and in the case of an incorrect answer what type of error was committed (i.e. logical, informational or statistical/arithmetic).

For the sample of N=1057 undergraduate medical students we report the mean number of correct responses on the progress test, stratified by year of training/education. For each of ChatGPT and GPT-4 we report the number/percentage of correct answers. We compare the performance of ChatGPT versus GPT-4 using McNemar’s test for dependent proportions. We report mean response times/lengths and compare these statistics between ChatGPT and GPT-4 using paired t-tests. We report the number/percentage of test questions for which ChatGPT and GPT-4 provided a rationale for the generated response, and compare these dependent proportions using McNemar’s test. A single physician (AP) with a background in medical education and test writing reviewed the responses of each AI chatbot, and labelled errors as logical, information, or statistical/arithmetic. We used counts/percentages to describe the types of errors made by each LLM-based chatbot (i.e. logical, informational, or statistical/arithmetic) [Gilson et al., 2023]. Logical errors occur when the chatbot identified relevant information to answer the question but did not convert this information into the correct answer. Information errors occur when the chatbot did not identify a correct piece of information (either from the question prompt, or general external information), that was needed to correctly answer the question. Statistical/arithmetic errors occur because of incorrect arithmetic by the chatbot.

The University of Toronto Research Ethics Board provided approval for conducting the study (REB-ID: 00044429).

## 3 Results

A total of N=1057 medical students from the University of Toronto completed an official undergraduate medical education progress test between Fall-2022 and Winter-2023. Student performance, in terms of mean number of correct responses, improved with increased training/education (Table 1): UME-Year1 mean=36.3%; UME-Year2 mean=44.1%; UME-Year3 mean=52.2%; UME-Year4 mean=58.5%.

**Table 1:** Performance of medical students (stratified by year of training/education) on the 100-item University of Toronto Undergraduate Medical Progress Test.

|  | Sample Size | Mean Correct | LL 95% CI | UL 95% CI | Minimum | Maximum |
| --- | --- | --- | --- | --- | --- | --- |
| UME-Year1 | 260 | 36.3 | 35.6 | 37.0 | 19 | 54 |
| UME-Year2 | 271 | 44.1 | 43.2 | 45.0 | 17 | 70 |
| UME-Year3 | 257 | 52.2 | 51.2 | 53.2 | 25 | 69 |
| UME-Year4 | 269 | 58.5 | 57.5 | 59.5 | 29 | 79 |

Comparatively, GPT-4 answered 79/100 (79.0%) questions correctly; whereas, ChatGPT answered 68/100 (68.0%) questions correctly. GPT-4 performed statistically significantly better than ChatGPT on the undergraduate medical education progress test (McNemar’s test: P=0.034). Remarkably, GPT-4 performed at a level equivalent to the top performing undergraduate medical student who attempted the examination.

In Table 2, we compare ChatGPT and GPT-4 with respect to response time, response length, rationale for answers provided and types of errors committed. We observed that mean response times for ChatGPT and GPT-4 were comparable (ChatGPT=15.3 seconds versus GPT-4=17.5 seconds: P=0.18). The mean response length for ChatGPT exceeded that of GPT-4 (ChatGPT=1517 characters versus GPT-4=1259 characters: P<0.0001). ChatGPT was more likely than GPT-4 to provide some type of rationale for the answer it provided (ChatGPT=98.0% versus GPT-4=84.0%: P=0.0005). And logical errors were more common than information errors (or statistical errors) for both LLM-based chatbots.

**Table 2:**
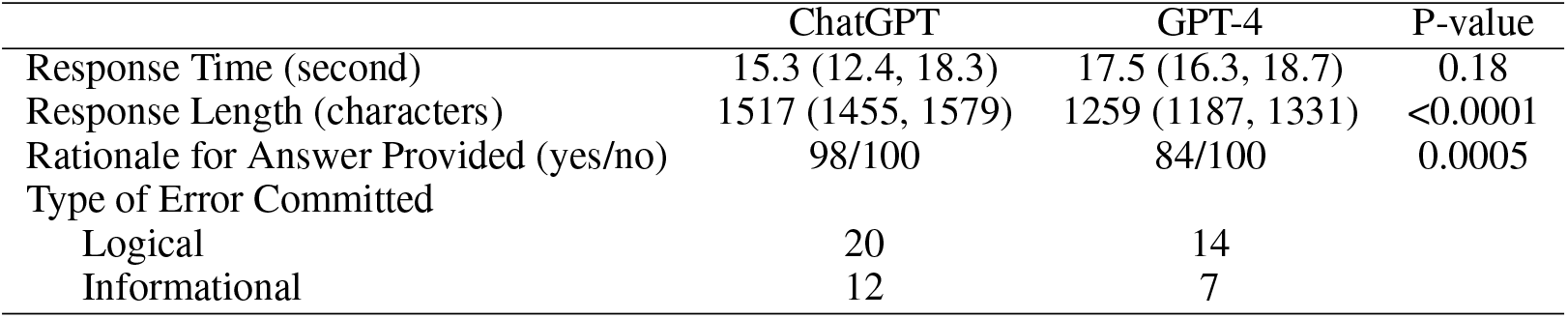
Response time, response length, rationale for answer provided, and types of errors committed by ChatGPT compared with GPT-4.

|  | ChatGPT | GPT-4 | P-value |
| --- | --- | --- | --- |
| Response Time (second) | 15.3 (12.4, 18.3) | 17.5 (16.3, 18.7) | 0.18 |
| Response Length (characters) | 1517 (1455, 1579) | 1259 (1187, 1331) | <0.0001 |
| Rationale for Answer Provided (yes/no) | 98/100 | 84/100 | 0.0005 |
| Type of Error Committed |  |  |  |
| Logical | 20 | 14 |  |
| Informational | 12 | 7 |  |

Response narratives for questions determined to be incorrect were reviewed for themes in processing. Three main themes were identified: 1) the AI missed or misinterpreted key features on history, physical examination or laboratory investigations required for the correct answer, 2) the AI did not appropriately prioritize critical investigation or management steps, or 3) the AI didn’t choose the most clinically practical or high value (Choosing Wisely) response. Table 3 provides an example of a progress test question and different responses between ChatGPT and GPT-4. The GPT-4 response includes a rational consideration of all potential responses and ultimate choice of the best response. The ChatGPT response considers the same information but fails to appropriately weight the risk for non-bacterial infection in this common clinical scenario (logical error).

**Table 3:** Sample progress test question with ChatGPT and GPT-4 responses.

| Progress Test Question | ChatGPT Response | GPT-4 Response |
| --- | --- | --- |
| A 69-year-old woman with acute myeloid leukemia is in hospital being treated for fever and neutropenia. She spiked her first temperature on day 7 of chemotherapy and continues to spike fevers of > 38.8 C despite 5 days of Piperacillin/ Tazobactam and Ceftazidime. She has no other symptoms. She has no lines. She was not previously on prophylactic antibiotics. She is hemodynamically stable. Her physical is unremarkable aside from a temperature of 38.8 Celcius. Her neutrophil count is still $0.4 \times 10^9/L$. Blood cultures have been repeated daily and are negative. A CXR is unremarkable. Which one of the following is the best next step?<br>A - Give a dose of Vancomycin<br>B - Add intravenous Itraconazole<br>C - Switch antibiotics to meropenem<br>D - Stop antibiotics and monitor | In a febrile neutropenic patient with persistent fever despite broad-spectrum antibiotic therapy, the best next step is to:<br>A - Give a dose of Vancomycin<br>Persistent fever in a neutropenic patient raises concern for resistant gram-positive organisms, such as methicillin-resistant Staphylococcus aureus (MRSA). Vancomycin is an appropriate choice to cover MRSA and should be added to the antibiotic regimen in this case.<br>The patient's clinical presentation, including persistent fevers despite 5 days of initial antibiotic therapy, neutropenia, and negative blood cultures, suggests the need for additional coverage. Vancomycin provides coverage against MRSA, which is an important pathogen in hospitals and healthcare-associated infections. It is important to consider adding vancomycin early in the management of febrile neutropenic patients who remain febrile despite initial broad-spectrum antibiotics.<br>Options B, C, and D are not the best next steps in this scenario. Adding intravenous Itraconazole (option B) is not indicated as there are no specific indications for antifungal therapy based on the given information. Switching antibiotics to meropenem (option C) is not necessary at this point since the patient has already been on broad-spectrum antibiotics (Piperacillin/ | This patient presents with persistent neutropenic fever despite broad-spectrum antibacterial therapy, which raises the concern for a fungal infection. Neutropenic fever is a medical emergency. The standard initial approach to neutropenic fever is to start broad-spectrum antibacterial agents immediately, targeting both gram-negative and gram-positive organisms.<br>However, if the patient has been on broad-spectrum antibiotics and continues to be febrile for several days, an infectious diseases consultation is typically obtained, and an antifungal agent is often added to the regimen. This is due to the concern for invasive fungal infections, which can be life-threatening in this population. Given the broad coverage of Piperacillin/ Tazobactam and Ceftazidime for bacterial pathogens, the next most likely source of infection in this setting would be fungal.<br>Therefore, the correct answer is:<br>B - Add intravenous Itraconazole.<br>Of note, Itraconazole is usually used in oral form for prophylaxis against fungal infections in immunocompromised individuals. Intravenous forms of other antifungal medications, such as Caspofungin or Liposomal Amphotericin B, are often used in the setting of suspected invasive fungal infections. However, the exact choice of antifungal agent would depend on several factors |

## 4 Discussion

In this study we compared the performance of ChatGPT and GPT-4 versus a cohort of medical students from the University of Toronto’s Temerty Faculty of Medicine on an undergraduate medical education progress test. A cohort of N=1057 undergraduate medical education students completed the 100-item multiple choice progress test between Fall-2022 and Winter-2023; and we observed performance increased with training level (UME-Year1 mean=36.3%; UME-Year2 mean=44.1%; UME-Year3 mean=52.2%; UME-Year4 mean=58.5%). Comparatively, ChatGPT answered 68/100 (68.0%) questions correctly, while GPT-4 answered 79/100 (79.0%) questions correctly. GPT-4 performed at a level equivalent to that of the top performing undergraduate medical education student who attempted the progress test (max score = 79/100).

The impressive performance of LLM chatbots on high stakes examinations across a variety of academic disciplines has been documented by OpenAI [2023]. Various authors have investigated the performance of LLM chatbots (particularly ChatGPT and GPT-4) in the context of high stakes medical examinations. For example, Gilson et al. [2023] investigated the performance of ChatGPT on the United States Medical Licensing Examination and demonstrated it was able to score >60%, which represented a passing score. Nori et al. [2023] extended these previous works, showing that GPT-4 performance exceeded that of ChatGPT when applied against the United States Medical Licensing Examination. A similar study was conducted by Kung et al. [2023], which validated the finding that ChatGPT was able to achieve a passing score on the United Stated Medical Licensing Examination. Kasai et al. [2023] evaluated the performance of ChatGPT and GPT-4 on the Japanese Medical Licensing Examination (2018-2022) and demonstrated that GPT-4 performance exceeded that of ChatGPT; and further, that GPT-4 was able to pass all versions of the licensing examination. Jang and Kim [2023] evaluated the performance of ChatGPT and GPT-4 on the Korean Medical Licensing Examination and demonstrated that GPT-4 performance exceeded that of ChatGPT; with GPT-4 nearly passing the examination. Thirunavukarasu et al. [2023] investigated the performance of ChatGPT on the Royal College of General Practitioners Applied Knowledge Test and demonstrated that it achieved a near passing score of 60%. Strong et al. [2023] evaluated the performance of ChatGPT on an open-ended free-text response clinical reasoning examination, and demonstrated that it was able to generate a passing response to nearly half of the questions posed. Huang et al. [2023] demonstrated that GPT-4 outperformed all residents who completed an official University of Toronto Family Medicine Residency Program Progress Test.

A growing body of literature is accumulating in medical education, demonstrating the remarkable performance of LLM-based chatbots on several high-stakes medical examinations. Without any specific fine-tuning on medically focused training datasets, these general-purpose artificial intelligence chatbots demonstrate expert/human-like knowledge of fact-based clinical reasoning. This has led researchers and medical educators to question how these artificial intelligence tools may be used to transform medical student learner experiences [Eysenbach et al., 2023]. Given the performance of LLM-based chatbots across a variety of medial examinations, Fleming et al. [2023] evaluated whether these artificial intelligence-based tools could be used to develop medical tests/examinations. More generally, Karabacak et al. [2023] propose opportunities of LLM-based chatbots in the creation of immersive learning environments which aim to enhance medical student learning opportunities through the creation of realistic (synthetic) clinical simulation scenarios, generation of digital patient documents, and through the provision of personalized feedback/evaluation. Further, Abd-Alrazaq et al. [2023] hypothesize that LLM-based chatbots offer opportunities to transform medical student/resident learning experiences and improve overall learner knowledge, skills and competence. The authors suggest that LLM-based chatbots might be used to develop entire medical curricula, augment existing teaching methodologies, generate personalized study plans and learning materials, and aid in student preparation of medical exams/assessments. Finally, Lee et al. [2023] investigated the potential benefits, risks, and opportunities of artificial intelligence based chatbots in medicine; as a tool for assisting/augmenting clinical practice, research and education. Future work should continue to investigate and evaluate how LLM-based chatbots might be integrated into medical education settings, to assist both medical learners and educators.

Our study is not without limitations. To begin, our study evaluated LLM chatbot performance on a single undergraduate medical exam, in the context of a single academic institution (University of Toronto). That said, this single institution study generated findings that were consistent with other similarly conducted studies, evaluating LLM chatbot performance on different medical examinations in different contexts. Additionally, in terms of evaluation, a single author/reviewer (AP) assessed LLM chatbot performance using the criteria laid out in Gilson et al. [2023]. Our study could be improved by employing multiple independent blinded reviewers/assessors and subsequently evaluating inter-rater agreement. Further, the evaluation criteria laid out in Gilson et al. [2023] are sensible and have face validity, hence why we chose to endorse them; however, future research should consider improved ways for qualitatively evaluating the responses generated by LLM chatbots with respect to medical education prompts. Finally, ChatGPT and GPT-4 are proprietary models (developed by OpenAI); other proprietary LLM’s have been developed by different technology companies. In the current social/legal context, the LLM training data, model architectures, computational hardware and optimization routines are considered proprietary and are generally unknown/non-transparent. Further, certain models only exist behind paywalls (e.g. GPT-4). These factors may limit the use of LLM chatbots in practical medical education settings.

## 5 Conclusions

This study adds to a growing body of literature demonstrating the remarkable performance of LLM-based chatbots on medical tests. GPT-4 performed at a level equivalent to the best performing undergraduate medical student, from a cohort of N=1057 medical students who attempted the official University of Toronto medical progress test in 2022/2023. Future work will investigate the application of LLM-chatbots as potentially transformative tools for learners/educators in University of Toronto medical education programs.

## Data Availability

University of Toronto Undergraduate Medical Education Progress Test questions (input prompts to ChatGPT/GPT-4) are considered proprietary information, and are not available publicly.

